# Short-term variability of proton density fat fraction in pancreas and liver assessed by multi-echo chemical-shift encoding-based MRI at 3 T

**DOI:** 10.1101/2021.06.08.21257560

**Authors:** Jürgen Machann, Maytee Hasenbalg, Julia Dienes, Robert Wagner, Arvid Köhn, Andreas L. Birkenfeld, Konstantin Nikolaou, Stephanie Kullmann, Fritz Schick, Martin Heni

## Abstract

**BACKGROUND:** Quantification of pancreatic fat (PF) and intrahepatic lipids (IHL) is of increasing interest in cross-sectional epidemiological and interventional studies in subjects at risk for metabolic diseases. Up to now, short- and medium-term variations as well as their dependence on actual nutritional status are almost unknown.

**PURPOSE or HYPOTHESIS:** To assess short-term intraday variations of PF/IHL after a high-fat meal as well as medium-term changes after 5 days of high-caloric diet with a 1500 kcal surplus on individual daily energy requirement.

**STUDY TYPE:** (retrospective/prospective/longitudinal/case control/cohort etc.) Prospective cohort study.

**SUBJECTS:** Twelve healthy subjects (6m/6f) for intraday variations, 15 healthy male subjects for medium-term high-caloric diet and 11 age- and BMI-matched controls.

**FIELDSTRENGTH/SEQUENCE:** 3 T whole-body imager (Magnetom Vida, Siemens Healthineers, Erlangen, Germany), assessment of proton density fat fraction by chemical-shift encoded MRI (multi-echo gradient echo sequence, qDixon).

**ASSESSMENT:** Manually drawn regions of interest in head, body and tail of pancreas as well as in liver by an experienced medical physicist carefully avoiding inclusion of surrounding visceral fat (pancreas) or blood-vessels (liver).

**STATISTICAL TESTS (please list the significance level):** Repeated measurements Anova for variabilities of PF and IHL, linear correlation analyses for relation of PF, IHL and BMI. Significance level p < 0.05 for all.

**RESULTS (must have numerical data and statistical testing for each phrase):** Non-significant changes in PF in both studies (2.5±0.9 vs. 2.5±1.0% after high-fat meal, 1.4±0.8 vs. 1.6±0.6% after high-caloric diet and 1.6±0.7 vs. 1.8±1.0% in the isocaloric control group), unchanged IHL after high-fat meal (2.5±0.9 vs. 2.4±1.0 %) and in the control group (1.1±0.6 vs. 1.2±1.1%), but significantly increased IHL after 5-day high-caloric diet (1.6±2.2% vs. 2.6±3.6%, p < 0.05).

**DATA CONCLUSION:** Daytime and nutritional status have no significant influence on ectopic fat depots in pancreas and liver and will therefore represent no major confounders in epidemiologic or clinical studies.

## Introduction

Due to the increasing prevalence of metabolic diseases – as type 2 diabetes mellitus – and cardiovascular diseases, there is a rising interest in detailed phenotyping of persons at risk. This is done during interventions, including lifestyle modification (1-4) as well as in epidemiological studies in the general population such as the Cooperative Health Research in the Augsburg Region (5), the German National Cohort (6) or the UK Biobank (7). For this purpose, magnetic resonance imaging (MRI) and proton magnetic resonance spectroscopy (^1^H-MRS) offer established and thorough techniques for quantification of whole-body adipose tissue distribution and assessment of ectopic fat deposition in abdominal organs as liver and pancreas, and in skeletal muscle. Adipose tissue volume and distribution throughout the body, as well as ectopic fat accumulation in parenchymal tissues are important markers for and presumably causative contributors to individual metabolic risk. In this context, pancreatic fat (PF) and intrahepatic lipids (IHL) are of major interest, as they are linked to impaired glucose metabolism. For PF, an inverse correlation with insulin secretion has been shown in persons with prediabetes (8-10) and it is hypothesized that PF modulates islet function in concert with further metabolic factors of the prediabetic milieu (11,12). Furthermore, a reduction of PF, e.g. by a dedicated lifestyle intervention, is thought to be pivotal for an improvement of pancreatic insulin secretion and the reversal of type 2 diabetes (13). Complementary, IHL are negatively correlated to insulin sensitivity (14). Indeed, non-alcoholic fatty liver disease (NAFLD) is the most common cause of chronic liver disease, progressing to inflammation and fibrosis (non-alcoholic steatohepatitis, NASH) (14), cirrhosis (15,16), liver failure and hepatocellular carcinoma (17). Even in normal weight subjects, fatty liver is a strong predictor for an unhealthy metabolism with increased risk for diabetes, cardiovascular events and mortality (18). IHL have therefore become one of the most important biomarkers in metabolic research.

Besides ^1^H-MRS, multi-echo gradient-echo chemical shift encoding-based techniques (CSE-MRI) have proven to be reliable for determination of proton density fat fraction (PDFF) (19-21) with some concomitant advantages. In contrast to ^1^H-MRS, the entire volume of an organ can be assessed within a short measurement time (<20 s, i.e. in a single breath-hold) with good spatial resolution, allowing a detailed analysis and intra-organ variabilities in fat-distribution. By contrast, when applying single-voxel ^1^H-MRS, information is obtained from a volume of several cm^3^. This is unproblematic in liver, as ectopic fat is evenly distributed, at least in subjects without liver pathologies (22). Repeated measurements, e.g., in the course of a lifestyle intervention, allow for a reliable assessment of changes in IHL when recorded at the same position. For the pancreas, this approach seems disputable.

Due to its lobulated form, its location, stretching from behind the stomach to the left upper abdomen, and its inhomogeneous composition (ducts, exocrine and endocrine tissues, inhomogeneous fat distribution), spectroscopic examinations of the pancreas are challenging and therefore in general not advised. Even small movements of the subject or minor organ movement/displacement between morphologic imaging and spectroscopic data acquisition may lead to inclusion of signal from nearby visceral adipose tissue. Therefore, imaging approaches are favored for quantification of PF.

Up to now, there is no information about short-term regulation of PF in the literature. Intra-day variations of ectopic lipids are described for intramyocellular lipids (IMCL), revealing large fluctuations in response to nutritional status and physical exercise (23), whereas IHL have shown to be almost inert in respect of fasting/fed state (24).

To clarify probable nutrition-induced variations in PF, we studied two cohorts of healthy subjects. The first underwent repeated CSE-MRI on the same day (two measurements in the early morning after overnight fasting and a third one after a high caloric meal) while the second was studied during the course of a 5-day high caloric diet with two examinations in the fasted state.

## Material and Methods

### Subjects

#### Study design

##### 1. Intraday variability

Twelve healthy subjects (6m/6f, age: 24-49 years, BMI: 19.2-27.8 kg/m^2^) were included in this part of the study. To test for reproducibility and intraday variability of PF and IHL, 3 examinations were performed (Fig. 1 A). First MR measurement was done in the early morning after overnight fasting, a second one within the same session after new adjustment of the scanner. The third examination followed at lunchtime, one hour after ingestion of an energy dense meal (pizza, containing 5100-7300 kJ, 52-93 g fat, 123-163 g carbohydrate) and a sweetened soft-drink (containing 53 g carbohydrate).

**Figure 1:**
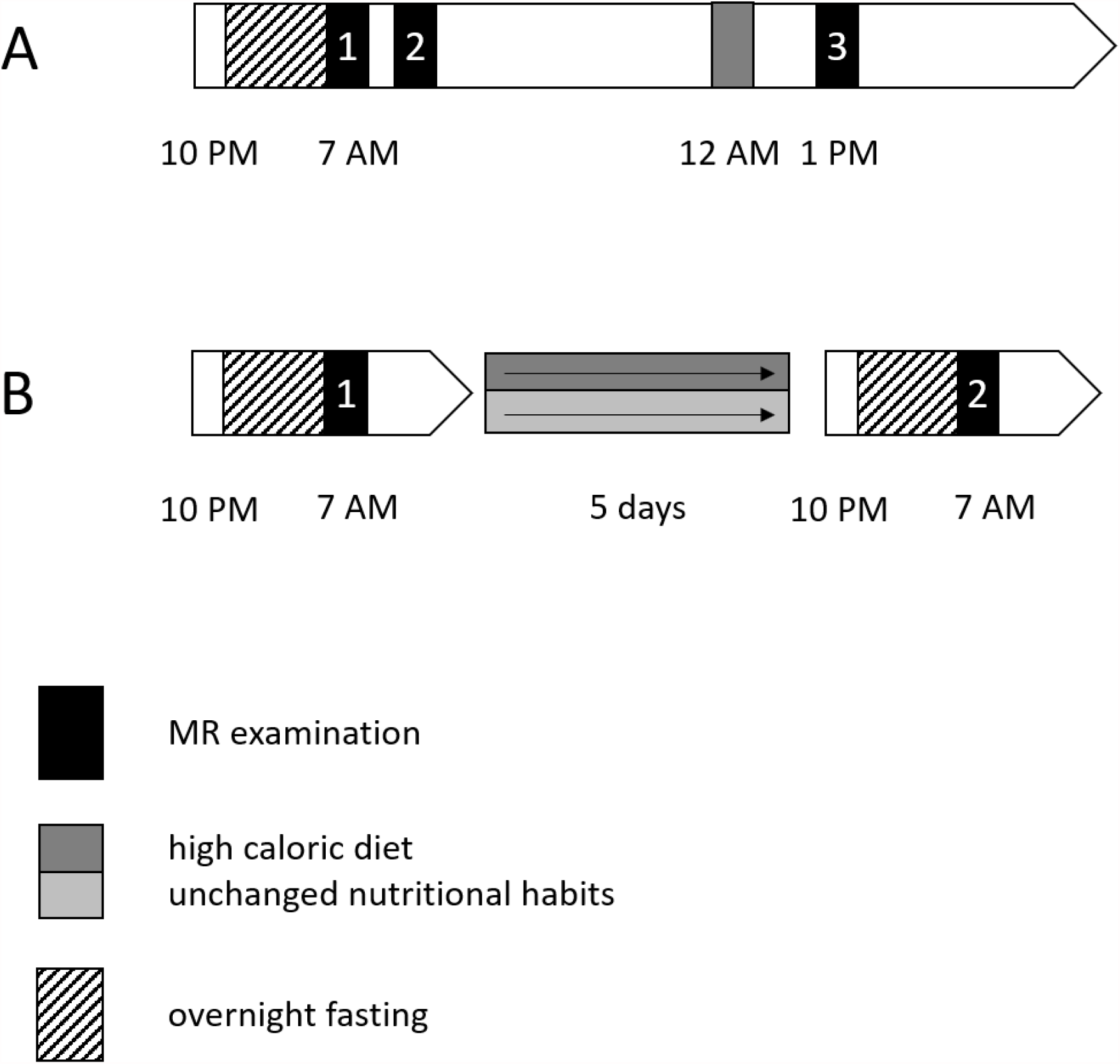
Flow-chart for repeated CSE-MRI in the framework of determination of intraday variabilities (A) and a 5-day high caloric diet (B)

##### 2. Variability after 5 days of high caloric diet

To assess medium-term changes, 15 healthy, young and lean male subjects (age: 19-27 years, BMI: 18.6-24.4 kg/m^2^) underwent a high-caloric diet for 5 days with a daily surplus of 6200 kJ on their individual basal metabolic rate. Additionally, 11 age- and BMI-matched males (age: 20-26 years, BMI: 19.9-24.0 kg/m^2^) were examined twice with the same time difference between the MR measurements and without changing their nutritional habits. MR examinations were performed in the early morning after an overnight fasting period at baseline and after the interval (Fig. 1 B). Both studies were approved by the local ethics committee and written informed consent was obtained from the volunteers.

#### MR examinations – study protocol

MR examinations were performed on a 3 T whole-body scanner (Magnetom Vida, Siemens Healthineers, Erlangen, Germany). Subjects were placed head first in supine position with the spine-array coil mounted on the patient table of the scanner. Additionally, an 18-channel body-array coil was used for homogeneous coverage of the upper abdomen. After morphologic imaging, a 3D multi-echo CSE sequence was placed to cover the entire liver and pancreas with following parameters: matrix size 160×132, field-of-view 380×314 mm, partition thickness 3 mm (80 partitions in total), repetition time TR = 8.9 ms, six echoes with echo times TE = 1.09, 2.46, 3.69, 4.92, 6.15 and 7.38 ms, flip angle 4°, acceleration Caipirinha, factor 2 in phase-encoding and slice encoding direction each, bandwidth 1080 Hz/pixel, 1 acquisition, acquisition time TA = 17 s (breath-hold). PDFF-maps were generated inline on the console of the scanner as described (20,25), correcting for microscopic magnetic field inhomogeneities by correction for T2*. In this PDFF-map, intensity values directly reflect PF and IHL in percent.

#### Post-processing

Quantification of PF and IHL was performed by manually drawing circular regions of interest (ROI) in the PDFF maps. Three small ROIs were selected in the pancreas, one in the head (PF_H_, Figure 2 A), one in the body (PF_B_) and one in the tail (PF_T_), both Figure 2B, carefully avoiding inclusion of surrounding visceral adipose tissue. The mean value of the three ROI’s was calculated (PF_mean_). Where necessary, evaluation of the three pancreatic subregions was done in different axial slices due to the lobulated form of the organ. Owing to the almost homogeneous distribution of fat in the liver, a larger VOI with a diameter of approx. 3 cm was chosen in the posterior part of Couinaud-segment 7 (see Fig. 2 C), carefully selecting the ROI in the identical position at the follow-up examinations.

**Figure 2:**
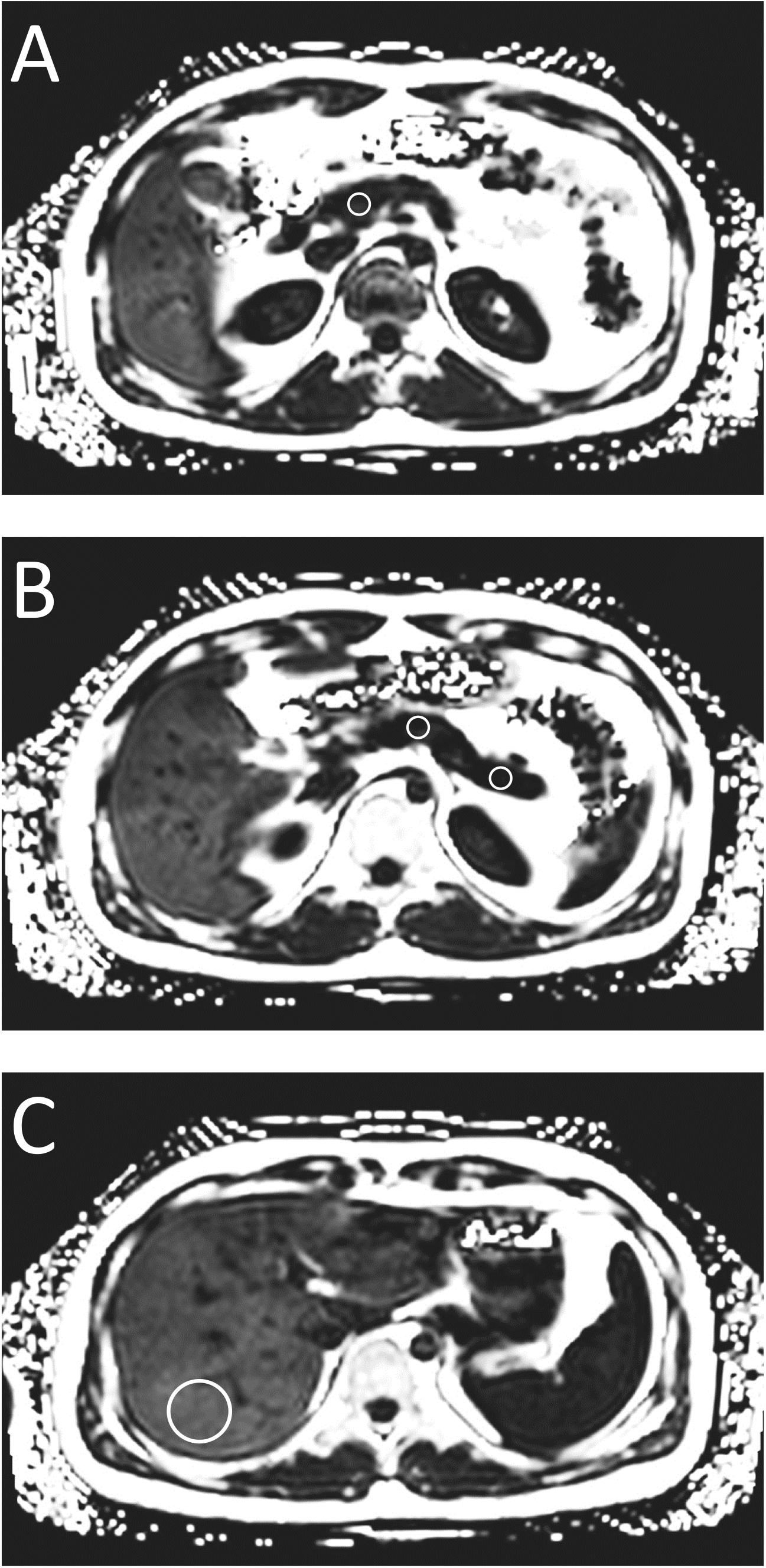
Axial PDFF-maps indicating ROI’s for evaluation of pancreatic proton density fat fraction (PF) in head (A), body and tail (B) as well as from intrahepatic lipids (IHL, C)

#### Statistics

Statistical analyses were performed with JMP (JMP® 15.2.0 SAS Institute, Cary, NC). Data are reported as mean ± SD unless otherwise stated. Evaluation of the effect of variabilities in ectopic fat compartments was done by a repeated measurement Anova analysis with a significance level set to p < 0.05. Univariate linear correlation analyses were used to analyze the coefficient of determination (R^2^) between PF, IHL and BMI. P-values<0.05 were considered statistically significant.

## Results

All MR examinations were of high quality and calculated PDFF-maps could be generated free of artifacts, thus being reliable for quantification of PF and IHL.

### 1. Intraday variability

Distribution of pancreatic fat in the twelve participating subjects revealed slight regional differences with PF_H_ being lowest (1.7±0.8%) whereas PF_B_ and PF_T_ were slightly higher (2.6±1.6 and 2.6±1.4%) in the first examination. Repeated measurement after new adjustment of the scanner resulted in almost similar PDFF values (1.7, 2.8 and 2.5% for PF_H_, PF_B_ and PF_T_, respectively). After the high-fat meal, insignificant variations were observed with mean values of PF_H_ = 1.7±0.7%, PF_B_ = 2.5±1.7%, PF_T_ = 2.8±1.3% (all p > 0.05 in a one-sided paired t-test). This finally resulted in PF_mean_ of 2.5±0.9%, 2.4±0.9% and 2.5±1.0% for examinations 1, 2 and 3 respectively. It has to be mentioned that the reproducibility in selection of the ROIs after ingestion of the meal was aggravated by relocation of the pancreas due to the fully loaded stomach as shown in Figure 3.

**Figure 3:**
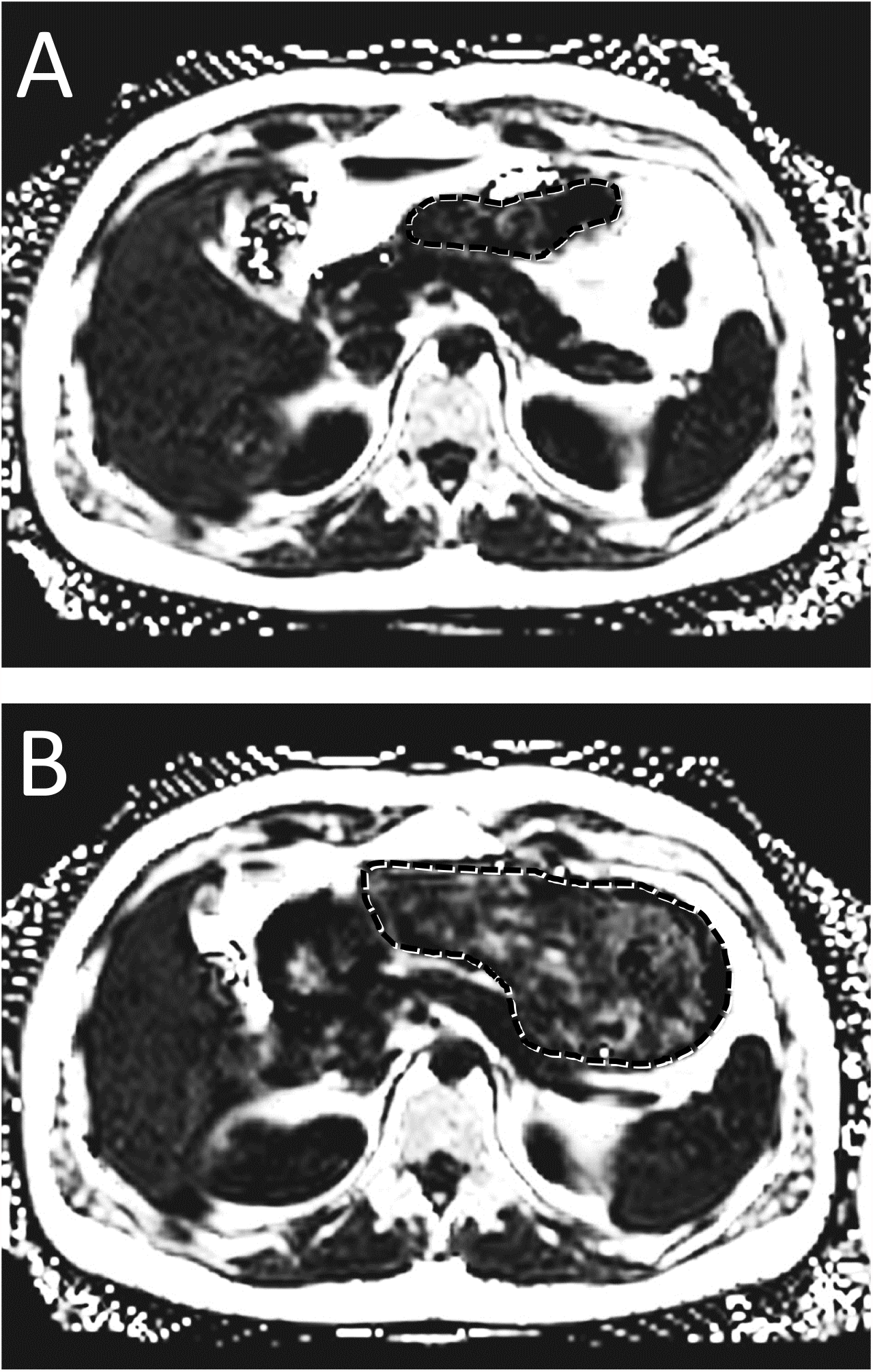
Axial PDFF-maps prior to (A) and after high fat meal (B) highlighting challenges of positioning the ROI due to the filled stomach (dashed line) at the second examination.

Individual progression of PF_mean_ is shown in Figure 4 A on the left side. There is a slight but insignificant increase of PF_mean_ in 8 subjects after the meal, 2 subjects remained unchanged and 2 slightly decreased PF_mean_.

**Figure 4:**
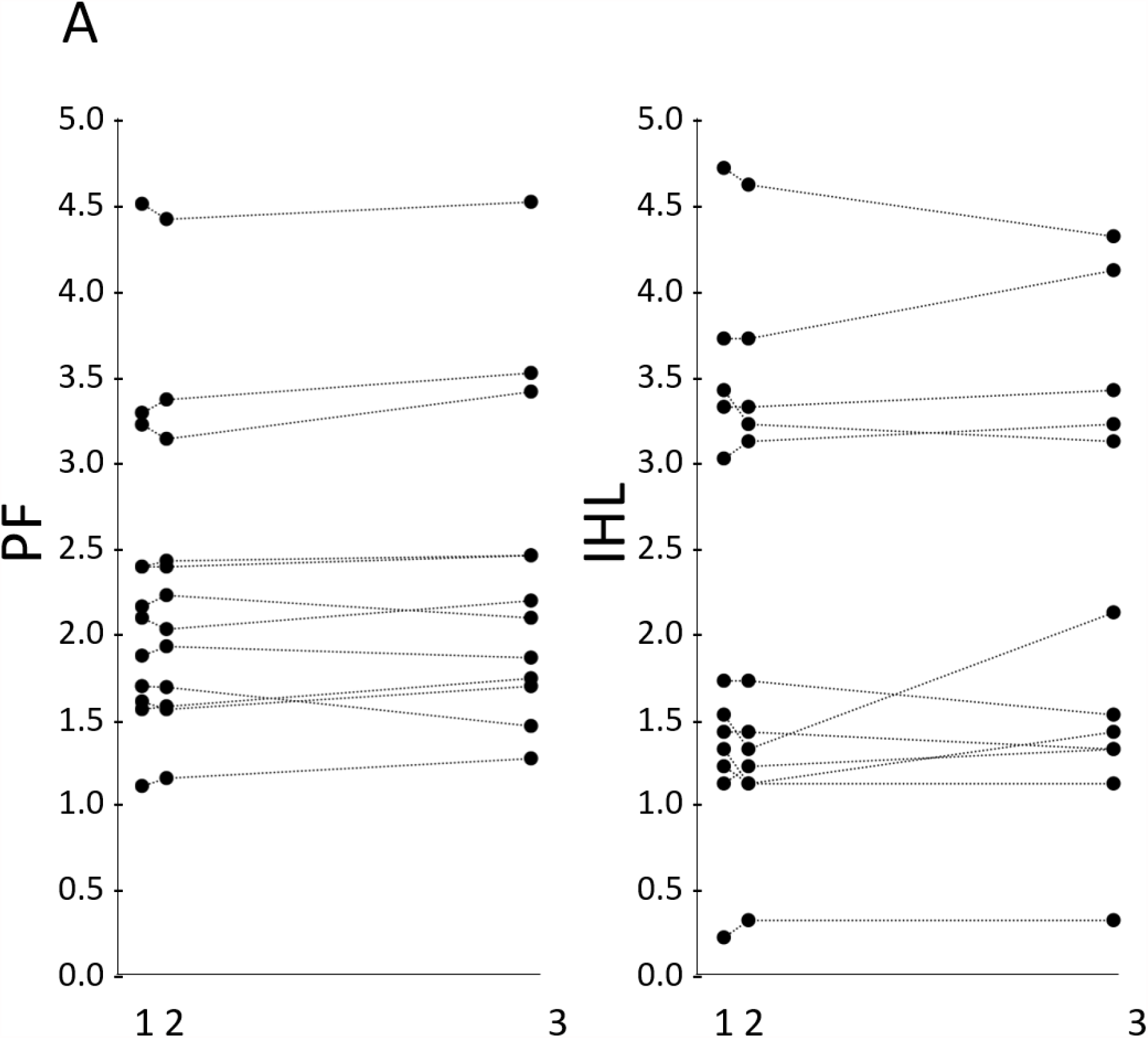
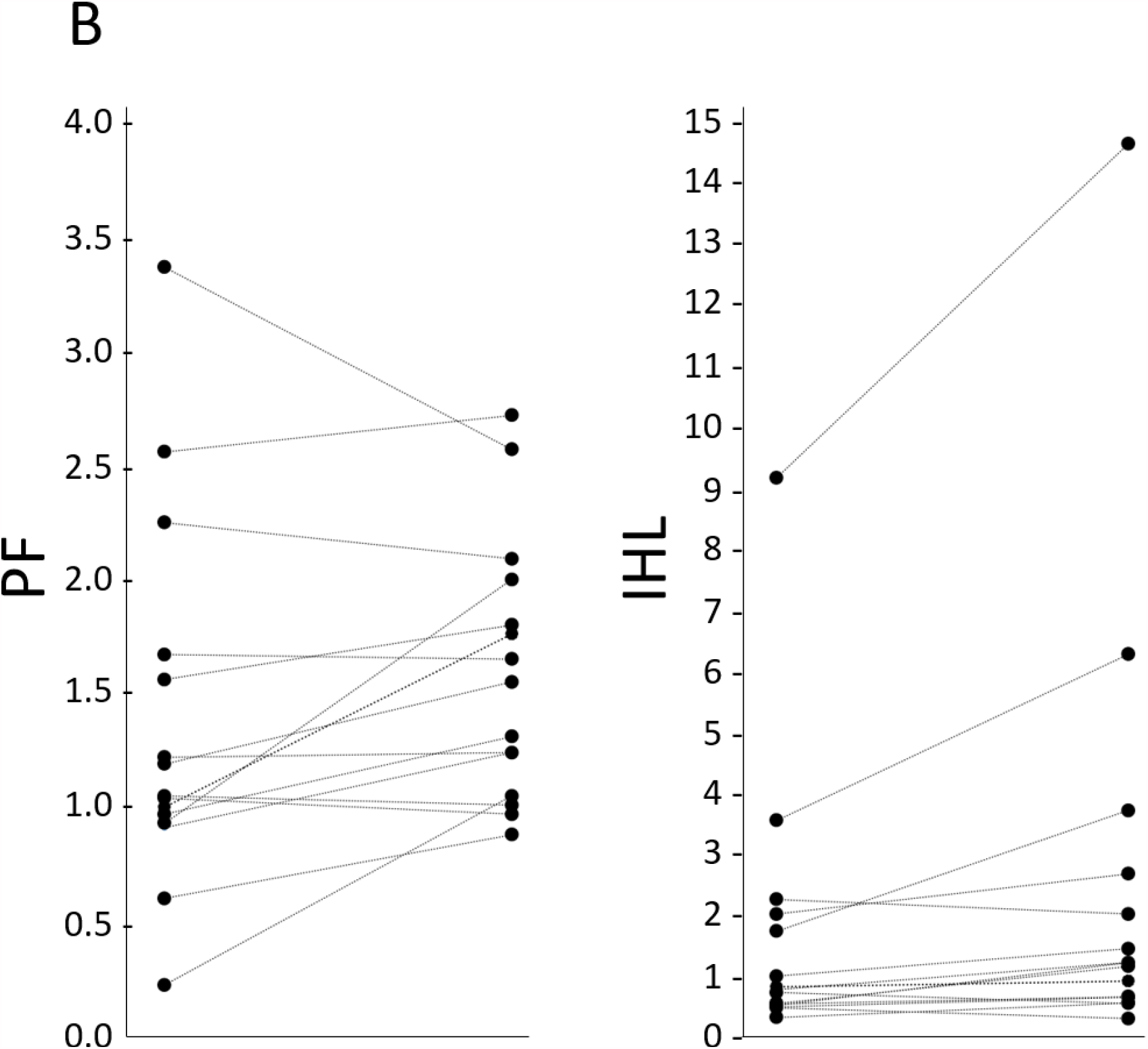
Individual variations of PF_mean_ (left) and IHL (right) in the course of repeated CSE-MRI. (A) intraday variabilities, (B) 5 days of high-caloric diet.

Almost assimilable results were obtained in the liver with IHL of 2.5±0.9% in the first measurement, 2.4±0.9% for the repeated measurement after new adjustment of the scanner and 2.4±1.0% one hour after the high fat meal (all p > 0.05). Regarding individual courses of IHL in Figure 4 B, a stronger increase of IHL was found in one subject (male, BMI 22.0 kg/m^2^) by 0.7%, corresponding to 50% from the baseline IHL content, whereas all other subjects remained almost unchanged or within the range of reproducibility. In total, IHL were increased in 7 and reduced in 5 subjects after the meal.

### 2. Variability after 5 days of high caloric diet

The regional distribution of PF in the 15 subjects participating in the 5-days high caloric diet revealed a comparable pattern with PF_H_/PF_B_/PF_T_ of 1.4±1.0%/1.3±0.9%/1.4±1.1% with PF_mean_ averaging to 1.4±0.8%. This pattern was slightly shifted after the dietary intervention resulting in 1.7±1.0% for PF_H_, 1.6±0.8% for PF_B_, and 1.5±0.9% for PF_T_, indicating a minimal mean increase in all subregions. Thus, PF_mean_ slightly increased from 1.4±0.8% at baseline to 1.6±0.6% after the high-caloric diet (n.s.). Individual courses for PF_mean_ are displayed (Figure 4 B, left side) showing a slight increase in 10 and a minimal decrease in 5 subjects.

Our observations of IHL showed in an increase from 1.6±2.2% (range: 0.3-9.2%) to 2.6±3.6% (range: 0.3-14.7%) reaching statistical significance in repeated measurement Anova analysis (p < 0.05). Within this, 12 subjects increased their IHL and 3 showed a marginal decrease as shown in Figure 4 B on the right side.

Within the control group of subjects not changing their dietary habits during this interval, a slight but insignificant increase in PF_mean_ from 1.6±0.7% to 1.8±1.0% (n.s., p = 0.54) was observed, IHL were unchanged (1.1±0.6% at baseline vs. 1.2±1.1%, n.s., p = 0.43).

There is a significant positive correlation between PF_mean_ and IHL in all subjects in both studies with an R^2^ of 0.16 and p < 0.05 as shown in Figure 5 A. Whereas PF does not show an association with BMI, IHL are significantly correlated with BMI with (R^2^ = 0.23, p < 0.01) as shown in Figures 5 B and C.

**Figure 5:**
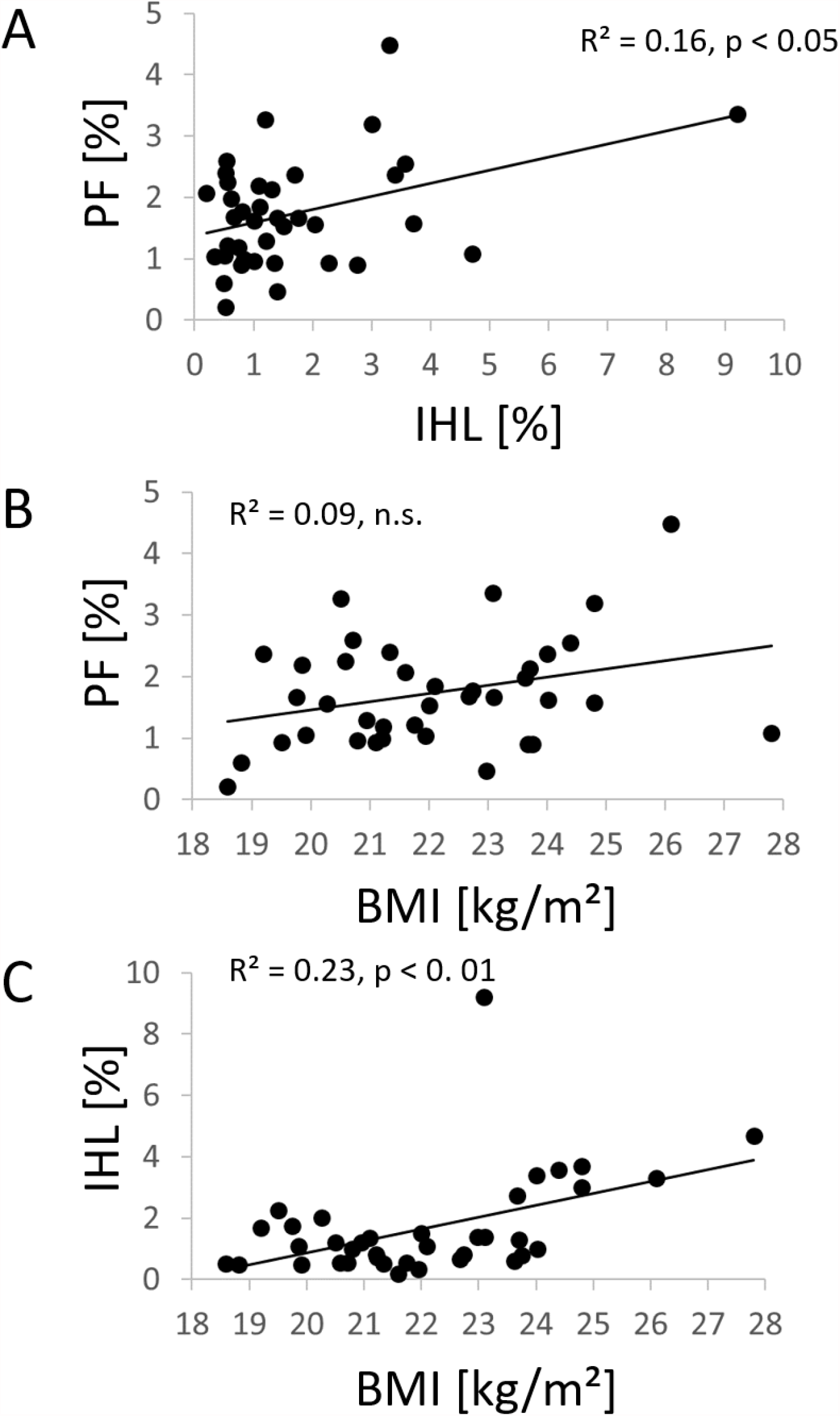
Linear regression between PF_mean_ and IHL (A), BMI and PF_mean_ (B) and BMI and IHL (C) reveal a positive but insignificant correlation between PF_mean_ and IHL and a significant positive correlation between BMI and PDFF in both organs.

## Discussion

Our data reveal only minor changes in PF as observed in three intraday measurements after a high fat meal as well as after 5 days of high caloric diet including a control group with unchanged nutritional habits. Hence, PF is a relatively inert ectopic fat depot. In contrast, IHL are significantly increased after the 5-day-dietary challenge, suggesting a more rapid accumulation of fat in the liver. Reproducibility of PF and IHL is shown to be very good in respect of intra-session results and the control group after 5 days of unchanged dietary habits.

Quantification of ectopic lipids, e.g. in skeletal muscle (IMCL), liver (IHL) or pancreas (PF) has gained a lot of interest in cross-sectional and longitudinal studies as ectopic lipid accumulation is a well-known crucial contributor to the pathogenesis of metabolic diseases (1,4,8-10,22,29). Thus, non-invasive MR-based phenotyping including fat quantification is increasingly being applied in large-scaled epidemiological studies as well as in interventional prospective studies. In contrast to the major adipose tissue compartments (subcutaneous fat and visceral fat), quantification of ectopic lipids in organs which – under healthy circumstances – contain little or no fat is more challenging and requires special techniques for exact assessment. Volume localized MRS has proven to be a reliable method, allowing a detailed analysis of characteristic metabolites in a specific tissue. E.g., IMCL, which have shown fast regulation in the course of short-term dietary intervention (26), fasting (27), exercise (28) or even in the course of the day (23) can exclusively be assessed by ^1^H-MRS. For quantification of IHL, ^1^H-MRS became the non-invasive gold standard (29) but is being replaced by CSE-based MRI meanwhile.

Due to the irregular lobulated shape and inhomogeneous fat distribution in the pancreas, ^1^H-MRS is not recommended for quantification of PF. Furthermore, availability of ^1^H-MRS is very limited whereas CSE-based MRI-techniques are increasingly obtainable. It has to be mentioned that two-point Dixon techniques are not suitable for this purpose as they do not offer the possibility to calculate PDFF.

CSE-based MRI is distinguished by excellent linearity, accuracy and reproducibility which has been shown in phantoms at different field strength and for different manufacturers (30-32) as well as for in-vivo applications (21,33,34). Thus, its application for quantification of PF is recommended. Whereas the acquisition strategy seems to be clarified, evaluation of the PDFF-maps is still debated. While the manual drawing of ROIs in pancreatic head, body and tail allows detection of regional variability of PF (9,35-37), this approach is prone to inter-observer discrepancies (38). The alternative strategy of volumetric (3D) analysis has improved repeatability and reproducibility (38,39). Al-Mrabeh et al. proposed a so-called “MR-opsy” approach, excluding signal contribution from pancreatic ducts and intrusions of visceral fat by thresholding and thereby excluding non-parenchymal tissue (40). In combination with automatic organ segmentation by deep-learning-based algorithms (41,42), this might promise a fast and user-independent post-processing procedure in the future.

Up to now – to the best of our knowledge – there is no information about short term variations of PF. Regarding long-term changes, a decrease of PF has been described after laparoscopic sleeve gastrectomy (43), after gastric bypass surgery in patients with type 2 diabetes mellitus, but not in subjects with normal glucose tolerance (44) and following diet-induced weight loss (45), but there is no information about changes in PF after high fat/high caloric diet in the literature. Long-term regulation of IHL after dietary lifestyle intervention is well known and described in the literature (e.g. 1,2,4,13,14,43) but little is known about the period when this reduction becomes evident.

The generalizability of our findings might be limited as we included only healthy and rather lean young subjects with relatively low PF and IHL; only one subject fulfilled criteria for fatty liver (i.e. IHL > 5.56 % (29)) who showed the strongest increase after the high caloric diet. Furthermore, we only evaluated two time-points in the intra-day study and can therefore not rule out variations at later time points.

As no significant alteration was observed in the intra-day examinations, confirming the results of Colgan et al. who studied 13 subjects with fatty liver (24), we conclude that there are no clinically relevant short-term/intraday variabilities, neither in the liver nor in the pancreas.

Another point which has to be addressed is, that this study was restricted to high caloric and high fat interventions. Whether or not there are short-term changes after prolonged starvation or after exercise as previously detected for IMCL (23) cannot be answered from our data.

When planning cross-sectional or longitudinal studies that aim to quantify PF or IHL, it is often discussed whether the MR examinations have to be performed in a standardized manner, e.g., in the early morning after overnight fasting or with a special dietary program in the days prior to measurements. Our current data indicate that daytime and nutritional status have no major impact.

## Data Availability

All requests for data will be promptly reviewed by the Data Access Steering Committee of the Institute of Diabetes and Metabolic Research, Tuebingen, to verify whether the request is subject to any intellectual property or confidentiality obligations. Individual-level data may be subject to confidentiality. Any data that can be shared will be released via a Material Transfer Agreement.

## Acknowledgments

We thank all study participants for their contribution and the Members of Siemens Healthineers for continuous support.

